# Fair machine learning models for disease prediction: In-depth interviews with key health experts

**DOI:** 10.1101/2025.02.04.25321632

**Authors:** Nhung Nghiem, Ramona Tiatia

## Abstract

Artificial intelligence (AI) and machine learning (ML) pose enormous potential for improving quality of life. It can also generate significant social, cultural and other unintended risks. We aimed to explore fairness concepts that can be applied in ML models for disease prediction from key health experts’ perspectives in an ethnically diverse high-income country. In-depth interviews with key experts in the health sector in Aotearoa New Zealand (NZ) were implemented between July and December 2022. We invited participants who are key leaders in their ethnic communities, including Māori (Indigenous), Pasifika and Asian. The interview questionnaire comprised six sections: (1) Existing attitudes to healthcare allocation; (2) Existing attitudes to data held at the general practitioner (GP) level; (3) Acceptable data to have at the GP level for disease prediction models; (4) Trade-offs for obtaining benefits vs generating unnecessary concern in deploying these models; (5) Reducing bias in risk prediction models; and (6) Including community consensus into disease prediction models for fair outcomes. The study shows that participants were strongly united in the view that ML models should not create or exacerbate inequities in healthcare due to biased data and unfair algorithms. An exploration of fairness concepts showed that carefully selected data types must be considered for predictive modelling and that trade-offs for obtaining benefits versus generating unnecessary concern produced conflicting opinions. The participants expressed high acceptability for using ML models but expressed deep concerns about inequity issues and how these models might affect the most vulnerable communities (such as Māori in middle-ages and above and those living in deprived communities). Our results could help inform the development of ML models that consider social impacts in an ethnically diverse society.

## Introduction

Cardiovascular diseases (CVD) and diabetes are among the top leading causes of death globally. (Wang, Naghavi et al. 2016) In Aotearoa New Zealand (NZ), diabetes and CVD are the leading causes of premature death and disease burden and major sources of health inequities for Māori (Indigenous), Pasifika, and Asian populations due to socio-economic, cultural and health system factors. These diseases are unequally distributed by ethnicity and deprivation, with Māori, Pasifika and groups with low socio-economic positions at a higher risk of obesity(Teng, Atkinson et al. 2016), CVD and diabetes.(Disney, Teng et al. 2017) The Covid-19 pandemic may have exacerbated these inequities.(Nghiem and Wilson 2021) Preventing CVD can be achieved with population-wide interventions (eg, tobacco control policies, pharmaceutical interventions and changes to the food environment eg, regulating sodium levels in processed foods).(Nghiem, Blakely et al. 2016, Nghiem, Cleghorn et al. 2018, Afshin, Sur et al. 2019, Nghiem, Knight et al. 2019, Nghiem, Leung et al. 2019) However, another strategy is to identify people at increased risk for better targeting more personalised lifestyles (diet, physical activity) and preventive pharmaceutical interventions.

Traditional models for CVD risk prediction, such as QRISK2, Framingham, and Reynolds, predict future risk of CVD based on well-established risk factors such as age, sex, high blood pressure, cholesterol, smoking, and diabetes.(Haq, Ramsay et al. 1999, D’Agostino 2008, Hippisley-Cox, Coupland et al. 2008, Riddell, Wells et al. 2010, Pylypchuk, Wells et al. 2021) However, there remains a large number of individuals at increased risk of CVD who fail to be identified by these models. In contrast, some individuals not at substantially increased risk are potentially given preventive treatment unnecessarily. (Brindle, Beswick et al. 2006, Ridker, Buring et al. 2007, Weng, Reps et al. 2017) Moreover, the prediction equations themselves may be sub-optimal due to missing or mismeasured predictors and may be due to the imperfect statistical model fit (e.g., the pre-specified relationship between a risk factor and outcome in (say) a Cox proportional hazards model). Improving this latter component, model fit is prone to over-fitting and is time-consuming; in this case, machine learning (ML) may assist with automated procedures to optimise prediction, including prediction harnessing of many different statistical model forms.(Athey and Imbens 2019)

Machine learning (ML) methods have proven promising new techniques for modelling risk prediction in a time of large datasets.(Crown 2015, Goldstein, Navar et al. 2016, Hofman, Sharma et al. 2017, Subrahmanian and Kumar 2017, Nghiem, Atkinson et al. 2023, Nguyen, Hy et al. 2023, Albrecht, Broderick et al. 2024, Nghiem, Wilson et al. 2024) ML methods consist of many alternative methods, including classification trees, random forests, neural networks, support vector machines, and lasso and ridge regression.(Kreatsoulas and Subramanian 2018) For studies where the primary goal is to predict the occurrence of an outcome, this technique produces a more flexible relationship between the predictor variables and the outcome.(Goldstein, Navar et al. 2016) It can accommodate non-linear relationships while overcoming the issues that overfit the traditional regression models.(Tay, Poh et al. 2015, Wolfson, Bandyopadhyay et al. 2015, Narain, Saxena et al. 2016) Established evidence suggests that ML significantly improves the accuracy of CVD risk prediction compared to traditional regression models.(Wolfson, Bandyopadhyay et al. 2015, Narain, Saxena et al. 2016, Weng, Reps et al. 2017, Barbieri, Mehta et al. 2021, Ravaut, Sadeghi et al. 2021, Nghiem, Atkinson et al. 2023, Nghiem, Wilson et al. 2024)

While a growing number of ML models have been developed to predict long-term health conditions, most of the models have not considered health inequity issues.(Cederholm, Eeg-Olofsson et al. 2008, Wang, Negishi et al. 2015, Kavakiotis, Tsave et al. 2017, Dagliati, Marini et al. 2018, Nguyen, Pham et al. 2019, Aminian, Zajichek et al. 2020, Fan, Zhang et al. 2020, Mehrabi, Morstatter et al. 2021, Ravaut, Sadeghi et al. 2021) A risk prediction model to predict CVD risks among people with diabetes has been developed in NZ.(Barbieri, Mehta et al. 2021) This risk prediction model used a deep learning approach and data for NZers aged 30–74 years with type 2 diabetes and without a history of CVDs. The factors of ethnicity (i.e., Māori, Pasifika, Indian, and Chinese or other Asian) were used as a prediction variable and did not provide any model evaluation by ethnicity. Another example is a CVD risk prediction for the Canadian population using administrative health data.(Ravaut, Sadeghi et al. 2021) That study used a wide range of socio-economic features in the ML prediction model, but no health equities were included in the model development.

Among the ML models in healthcare that included fairness concepts, mathematical constraints were used to impose fairness, such as narrowing gaps in model performance by ethnicity.(Zink and Rose 2020, Diana, Gill et al. 2021, McGuire, Zink et al. 2021, Nghiem, Atkinson et al. 2023) To avoid unfairness due to misclassification rates (such as higher wrong prediction rates for women than for men), Zafar et al. (Zafar, Valera et al. 2017, Zafar, Valera et al. 2017) introduced intuitive measures of disparate mistreatment for binary decision boundary-based classifiers (e.g., logistic regression). They showed that the method effectively avoids disparate mistreatment with a small cost in terms of accuracy. This fairness method was extended by Zink and Rose (Zink and Rose 2020) for continuous outcomes, i.e., health care payments, to provide fair benefits and health care coverage for all enrollees, regardless of their health status. Using both real (the IBM MarketScan Research Databases) and simulated data, they suggested that their new fair regression methods may lead to massive improvements in group fairness with only small reductions in overall fit.

A few studies have attempted to include participants’ perspectives of local communities in _ML_ models.(Chouldechova, Benavides-Prado et al. 2018, Kawakami, Sivaraman et al. 2022, Kuo, Shen et al. 2023) The case study of an algorithm-assisted decision-making programme in child maltreatment involved the interviews of impacted families in various stages of the ML model development process.(Chouldechova, Benavides-Prado et al. 2018) There is a growing number of studies investigating human-centric ML models,(Ho and Wang 2021, Kawakami, Sivaraman et al. 2022) However, these studies have primarily focused on modelling or policy discussion without involving the communities. To our knowledge, no studies have attempted to interview key health experts to inform fairness in CVD and diabetes prediction ML models. Given this background, we aimed to explore fairness concepts that can be applied in ML models for disease prediction from key health experts’ perspectives in an ethnically diverse high-income country: NZ.

## Methods

In-depth interviews with key experts in the health sector in NZ were implemented between July and December 2022. We invited participants who are key leaders in their ethnic communities, including Māori (Indigenous), Pasifika and Asian.

### Participant recruitment

Participants for the study were recruited from across the North and South Islands of Aotearoa/NZ for their knowledge and understanding of NZ health system outcomes on the lives of Māori, Pasifika and Asian ethnic communities. The researchers recruited various contact networks, and the snowballing process found a small number. In the two weeks before the day of the interview, participants were sent information and consent forms for the study to be recorded and transcribed. Nine participants, in total, were interviewed online using Microsoft Teams and were also informed that they were free to withdraw from the study at any time. The study concluded at the end of March 2023.

### The questionnaire design

We aimed to present participants with a range of health and social data sources that could be used to develop ML approaches to diabetes treatment and prevention (but which may also be relevant to many other diseases). We also wanted to investigate how participants defined “fairness” and explore how predictive models can prevent or avoid discriminatory health outcomes against any particular group of NZers.

The interview questionnaire has six sections. The week before the research team met with a participant, a copy of Section IV (trade-off scenarios) was sent so the participant could have time to read and consider the scenarios. This reduced some of the reading material provided during the interview.

At the beginning of the interview, participants were presented with open-ended questions in *Sections I* (existing attitudes to healthcare allocation) and *Section II* (existing attitudes to data held at the general practitioner level). Open-ended questions allowed the participants to speak freely about their experiences within the health system and whether they handled and used different data sources in their professional work. *Section III* (acceptable data to have at the GP level for disease prediction models) contains an itemised list of data source types in which participants were asked to assess a range of government administrative data. Participants were asked to go through the list with the hypothetical scenario in mind: if this was information about them or someone they knew, how comfortable would they feel if it were made available for predictive modelling? Once participants finished rating themselves with a score between 1 and 5 the researchers spent a short time asking them to give their reasons behind their scores.

In *Section IV* of the study, the participants were then provided with three *Scenarios of Trade-offs* for obtaining benefits versus generating unnecessary concern to explore variations of meaning associated with fairness, such as harms and benefits. Participants were asked to indicate whether they agreed or disagreed with the posited trade-offs.

*Section V* explored *Reducing bias in risk prediction models*, where participants were asked whether they agreed or disagreed with the proposed statements; and *Section VI* examined methods of community consensus and participants’ views of these being used to design disease prediction models.

### The interview method

Two team members conducted the interviews with a consistent format for all participants. Each participant was interviewed on their own by the research team. The interview started with a warm word of welcome to the participant, self-introductions of everyone present, and an overview of the study. Following the order of the questionnaire, one team member read each interview question aloud, giving the participants time to clarify anything they wanted. The second team member took responsibility for making manual notes during the interview and asked for the participants’ permission to record the interview. The interview ran for approximately one hour, and after the interview, participants were offered a koha of a gift voucher worth NZ$50 (US$32) to thank them for their time and attendance. This is not a payment but a koha/gift of appreciation, which the participants do not have to pay back. The participant’s identities were kept confidential, and their responses were anonymised. We did not expect any direct benefits to the participants personally from completing this study; however, the information gathered will benefit the participants’ communities by making health systems fairer. We were unaware of any risks for the participants to participate in our research. Eight interviews were conducted via video conference, and one was conducted in person (but was recorded by phone). Of note was that the interviews were implemented during the Covid-19 Omicron outbreaks in NZ.

### Ethics approval

The study was approved by the University of Otago ethics approval processes, reference numbers HD20/012 and D22/101. No patients were directly involved in this study.

### Data analysis

The interviews were recorded and transcribed into text. The transcripts were first imported, coded and analysed using the NVivo software, with each question as a code. Next we created a list of quotations of each question for each participant in Excel to summarise our thoughts and findings. The close-ended response for each question was coded in Excel. All close-ended questions were calculated for count and mean values, value ranges and percentages. Lastly, the research findings were formatted in the order of questions in the questionnaire.

## Results

### Overall

Nine participants with health and data backgrounds, both males and females, from Māori, Pasifika and Asian communities were interviewed. The participants’ professional occupations included academics, statisticians, data managers, medical practitioners, and government and private industry employees. Most key leaders stated that health data are not currently used to allocate health care to address needs fairly. Participants indicated high levels of discomfort related to lifestyle data such as financial spending, food and diet, alcohol consumption, and employment. There was also a consensus view regarding the lack of useable data that are generally held in primary health care settings and whether it is acceptable for government agencies to share data on individuals (e.g., social welfare data) that would allow general practitioners (GPs) to create GP-level disease prediction models as part of its business model for example service delivery. While the trade-offs for obtaining benefits versus generating unnecessary concern also produced conflicting opinions, the ML models were highly accepted for predictive purposes. However, participants expressed concerns regarding inequity issues and the solutions that are needed to prevent the most vulnerable communities (such as Māori in middle-ages and above and those living in deprived communities) from being determinately impacted. A descriptive analysis of the survey results is presented in Table 1. Presented below are the key sections of the questionnaire the participants were asked, and their responses.

**Table 1.** Summary of the participants’ responses to close-ended questions in the questionnaire.

| Questions asked in the one-hour interview | No (%) | Yes (%) | Range |
| --- | --- | --- | --- |
| 1a_Existing attitudes: fairly allocated health system | 100 | 0 | Yes/No |
| 1b_Existing attitudes: GPs have enough individual-level data to make decisions | 89 | 11 | Yes/No |
| 2_Is this ok for the government to give individual data to GPs for prediction models | 22 | 78 | Yes/No |
| 3a_See scores at the bottom of this table | NA | NA | NA |
| 3b_Additional information can be given to GPs | 0.0 | 100.0 | Yes/No |
| 3c_Any information that should not be given to GPs | 22 | 78 | Yes/No |
| 4a_Tradeoffs Scenario 1: 1 slightly harm to 10 benefits extra 6 months of life | 89 | 11 | Yes/No |
| 4b_Tradeoffs Scenario 2: 5 or 10 harm to 10 benefits 6 months of life | 56 | 44 | Yes/No |
| 4c_Tradeoffs Scenario 3: what is your comfortable ratio harm to benefit (Of note was that the responses were very wide in value ranges, so we did not calculate the median ratio) | 22 | 78 | Yes/No |
| 5a_Reducing bias: ML models should not be used until it's guaranteed to be fair | 44 | 56 | Yes/No |
| 5b_Reducing bias: ML should be used if a slightly unfair but improved prediction | 22 | 78 | Yes/No |
| 5c_Reducing bias: What is fair in your view | 11 | 89 | Yes/No |
| 6a_Including community consensus: holding a hui | 11 | 89 | Yes/No |
| 6b_Including community consensus: polis system | 33 | 67 | Yes/No |
|  | Mean score on a 5-point scale |  |  |
| 3a_Demographic | 4.2 |  | 3 - 5 |
| 3a_Health_Health history | 4.3 |  | 1 - 5 |
| 3a_Health_The amount of alcohol | 3.3 |  | 1 - 5 |
| 3a_Health_Smokes tobacco | 4.1 |  | 1 - 5 |
| 3a_Health_Travels | 4.1 |  | 1 - 5 |
| 3a_Health_Person's diet | 3.6 |  | 2 - 5 |
| 3a_Health_Physical activity levels | 4.3 |  | 2 - 5 |
| 3a_Socio-economic_Immigration | 4.2 |  | 3 - 5 |
| 3a_Socio-economic_Spending | 2.3 |  | 0 - 5 |
| 3a_Socio-economic_Employment, occupation | 3.4 |  | 1 - 5 |
| 3a_Justice | 3.1 |  | 1 - 5 |
| 3a_All data | 2.7 |  | 0 - 5 |

Notes: “Yes and No”/“Unsure” re-classified as “No”;

#### (1) Existing attitudes to healthcare allocation

The first question was whether participants felt healthcare resources were allocated fairly in NZ. The results showed that all participants (n=9, 100%) think that health data in NZ is not currently used to allocate health care fairly to address communities’ treatment and prevention needs.

Most participants (89%) did not think GPs typically have enough individual patient data to base the best health decisions for individual-level care, while 11% thought otherwise. Based on their experiences of managing GP level data and other available data sources participants highlighted dense nature of unstructured formats (e.g., text and images), and the great difficulty of deriving value or utilisation when data quality was poor or inaccessible.

One participant suggested that to address the ongoing disparity, every part of the system needed to take into account data of population groups at the highest risk level.

Health data can offer great insights about the challenges an individual and their community are exposed to. However, as another participant stated, what often happens is that information has not been effectively translated into system changes. Despite numerous health innovations, there has been minimal progress over the last 50 to 60 years, especially for Māori.

Having explored the participants’ observations of healthcare allocation, we then sought their views about how data sources might be used for predictive modelling within a healthcare setting, namely, primary health general practice services.

#### (2) Existing attitudes to data held at the general practitioner (GP) level

When asked about the types of data they were aware of held by GP services, most participants (78%) felt confident that government administrative data (e.g., social welfare data) should be made available to GPs to improve how GP-level disease prediction models may work, as long as there were safe and transparent processes for doing so.

One participant said that information about an individual’s culture, including their history and food habits, are important factors a GP uses to formulate a treatment plan for a patient. The absence and exclusion of various social and cultural contextual elements can however, also lead to unfair treatment of a patient rendering a less holistic presentation of the patient, and this impacts the quality of predictive modelling.

Participants offered other reasons why they felt health care is unfairly allocated: (a) data is incomplete or missing because some people never go to a doctor or miss appointments, thus creating gaps about the true numbers about a residential area or ethnic community requiring health assistance at any point of time (b) patient interactions with their doctors is influenced by a doctor’s social background and affects the extent to which they can fully communicate with Māori, non-Māori and English speaking patients; (c) medical curriculums can often not prepare GPs about cross-cultural issues or how to work in Māori landscapes; (f) GP services have inadequate resources for collecting, processing and auditing information data sources, partly because of staffing shortages but also due to other high demand for services.

Participants reported that the COVID-19 pandemic has also exacerbated challenges of data utilisation. There was wide acceptance that safe data haven and co-governance processes are important to have in place to ensure that data is checked and shared for their intended purposes. Those who people spoke about shared data environments felt co-ownership and co-governance mechanisms would greatly benefit communities and individuals. Differences in how data collection processes among GP services are conducted around the country was identified as significant gap and that there was major need for standardisation. In addition to GPs day-to-day client caseloads, it was mentioned that referral links could be improved to government wrap-around disability, housing and social services for example, so that greater numbers of patients requiring care packages could be better assisted.

In this section, participants opened up immediately about their knowledge of health data and the realities of working with patchy and low-quality data, poor data linkage and the kinds of opportunities they saw to improve and develop better solutions. Currently, there are many administrative obstacles within GP services to prevent potential misuses of data. In the next section, we explored at a much more granular level the sharing and accessibility of data owned by the participants themselves and what expectations they in how their data should be handled and used within a primary care setting. These are presented in greater detail in the following section.

#### (3) Acceptable data to have at the GP level for disease prediction models

In the section three of the questionnaire, participants were asked to consider how comfortable they would feel about their private information being provided to their local primary healthcare provider by external sources (i.e. government agencies). They were then given the scenario, where personal information, formatted as official government administrative data was distributed and used by their GP primary care service for the express purpose of conducting health prediction modelling to improve day-to-day patient case management.

Table 1 (#3a) provides the full list of data information that are being shared by government sources to GP services the participants were asked to consider and then rate using a score between 1 and 5 to show levels of self-comfortability (5-point scoring scale: Very comfortable [5], to Very uncomfortable [1]). After the participants provided their scores, they were asked to provide comments about what led them make their decisions, and these were also collated.

As shown above, the lowest mean scores were Socio-economic Spending (2.3%), and All Data (2.7) were information that participants felt *most uncomfortable* to have shared with their GP. Justice (3.1), Alcohol Consumption (3.3), Employment (3.4), and Dietary Intake (3.6) follow closely in order at the mid-range level of comfortability/uncomfortability. Interestingly, these six datasets, had generated the greatest number of comments about data quality and end-user utilization. Participants shared concerns associated with data privacy, end-user biases, ethnic and racial profiling and stereotyping. Most of the participants held similar views and concerns about individual behavioural lifestyle factors, associated with food, dietary factors and alcohol consumption, can often draw negative attention to Māori and Pasifika communities. Participants specifically acknowledged how obesity and diabetes complications are most prevalent for these communities. A significant finding which elicited *the greatest concern and criticism* by participants was data associated with *financial spending.* This issue was by far the most *important data* that participants said should never because personal finances and how one spends their money is the private business of individuals. Money, like food and alcohol use, and lifestyle decisions were largely misunderstood because of cross-cultural and unconscious biases. Participants said that they observed multiple settings where these data were often misconstrued or devalued. Rather than being included as holistic lifestyle factors linked to an individual’s social capital, such as collective food rituals, indigenous protocols, cultural identities and ethnic languages. Lifestyle and behavioural factors, particularly financial spending on food and leisure activities are often used negatively to stigmatise and unfairly judge indigenous and ethnic minorities.

By contrast, Health History (4.3%), Physical Activity (4.3), Demographic (4.2%), Immigration (4.2), Tobacco Use (4.1), and Mode of Travel (4.1) were data that participants were less uncomfortable and could shared more readily to GPs than other data. Generally, it was agreed that because these data are protective factors of well-being, there would be useful inclusion of these for predictive modelling. Tobacco, was another topic participants strongly agreed as data that can be shared because of its evidentiary harmful impacts to health. As highlighted above, participants felt more concern and uncomfortable about individual data associated to alcohol use and money. But felt greater comfort levels in relation to tobacco, health history, physical activity, demographics, immigration and mode of travel where the benefits of data-sharing were widely known.

One participant said that given NZ’s ethnically diverse population, Eurocentric frameworks that underpin healthcare systems are failing to accommodate multi-generational households in Māori and Pasifika communities. Educational achievements and Justice were also often negatively misconstrued across many governmental settings. Participants felt that serious caution was needed especially by data end-users who hold little knowledge and understanding, or may be apathetic to the severe mortality rates and comorbidities experienced within these communities. As reiterated by on participant, important contextual factors such as engagement levels in school, the possibility of Māori leaving high school early to enter the labour force and impacts of institutional racism must all genuinely be considered in order to create inclusive and equitable data frameworks, like predictive learning models.

Another participant shared that despite the potential benefits of ML in healthcare, NZ’s data systems are far off from being able to support ML and AI innovations. This comment was made specifically with respect to the Integrated Data Infrastructure (IDI) that links individual-level data held by the NZ government (Stats NZ 2019). For them, it highlights the lack of the necessary computing power and data processing capabilities that the IDI systems currently offer and that there are far better options that should be explored for investment.

Many also participants felt concerned that the current model of bringing data together poses multiple risks, such as data privacy and security, and high costs of technological and information data. This in turn, leads to greater vulnerability for mistakes and misuses, as well as greater levels of mistrust in the government’s ability to protect, store, and encrypt private information held about its country’s citizens.

As one participant stated, individuals should have control over their own data, including knowing who has access to it and what it is being used for. All participants raised the concept of data sovereignty as being very important for ensuring that Indigenous communities have control over both the management and use of data of which they have sovereign ownership.

Concerns about combining and sharing data sets and the risks associated with utilisation can and does already perpetuate existing inequities for marginalised communities in Aotearoa, a grave concern that participants of the study felt needed to be seriously addressed. By encouraging communities to be involved in co-designing and developing prediction models, health researchers and data managers need to take responsibility for explaining what technical concepts are involved by using plain language. Translating these will help to build relevant solutions by which ethnic communities primarily should be able to benefit from, an issue which is discussed with participants later in section 6.

#### (4) Trade-offs for obtaining benefits vs generating unnecessary concern in deploying ML models

Table 1, 4a, 4b and 4c provide the results for three Trade-off Scenarios that were presented to the study participants who were asked to respond with: ‘agreed’, ‘disagreed’ or ‘not sure’.

##### The Trade-off Scenario 1

<u>One</u> patient gets slight health benefits from better GP-level prediction – but <u>one</u> is made slightly worse off because of being told they are at risk of an adverse health outcome when this is not actually correct. For example, one extra person with diabetes is put on blood pressure medicine that will help prevent heart attacks, but one extra person is put on these medicines unnecessarily. All those who take the medicine gain an average of 6 months of extra life; while the person taking medicine unnecessarily has mild side-effects – some slight extra tiredness and has to visit their GP two times extra per year. The majority of the participants (89%) disagreed and 11% agreed with this trade-off.

##### Trade-off Scenario 2

posited that <u>five to 10</u> patients get slight health benefits from better GP-level prediction – but <u>one</u> is made slightly worse off because of being told they are at risk of an adverse health outcome when this is not actually correct. Most (56%) still did not favour this trade-off, although 44% did support it.

##### Trade-off Scenario 3

Participants were asked to state what they deemed to be an accepted ratio of harm to benefit given any trade-off scenario. Although the individual ratios are not listed in Table 1, only 78% of participants provided a response. The remaining 22% felt too uncomfortable to give any number due to their belief that no one should ever be harmed.

When asked to explain why they supported the tradeoff scenarios, one participant said it was not feasible to ensure complete fairness in data analysis.

Others said it was essential to continue making a conscious effort to evaluate and address any potential biases. In this context, transparent fairness assessment is a solution that most participants felt was a necessary and standardised requirement that should be rigorously applied to predictive modelling procedures.

A few participants strongly recommended that all disease prediction models undergo routine, rigorous, and equitable assessments. Neglecting this crucial step would result in poor-quality data.

One participant noted that it was difficult to give a definitive answer about any of the trade-off scenarios because they largely depended on individual life circumstances, many of which are unknown by treating physicians or by the patients themselves.

Other participants felt that if the person receiving the benefit was young and the person who had to go to the GP a few extra times was older, then a life course perspective could be applied to this type of scenario to help with modelling.

Another participant insisted that there are always multiple aspects with trade-offs that can create ethical, legal and social complications or that these reappear as “colonialism all over again”. As an example, although a 10 per cent improvement in model prediction is conceivably still better than no improvement at all, the impacts and inequities for disparate indigenous and minority groups needs to be addressed as at the macro and local level. In this sense, it is crucial to proactively seek solutions to make predictive models work better for everyone.

In summary, the statements about trade-offs between obtaining benefits and generating unnecessary concern in deploying ML models produced diverse responses, with many participants agreeing that assessing harms and benefits against individual versus public health gains and losses is extremely important.

#### (5) Reducing bias in risk prediction models

In this section, the participants were asked two questions to examine levels of bias permitted in an ML model.

***Statement 1***: If disease risk prediction models are not well designed then they can work better for some groups in society rather than others (i.e. usually benefiting the groups which are largest in society). Yes or No? Reponses were fairly even, although more participants answered No (56%) than Yes (44%).

***Statement 2***: The participants were asked about their view on the issue of disease prediction using ML not being used in NZ until it is guaranteed to be fair (performing equally well for all groups and not better for one group than another). Yes or No? Three-quarters of participants (78%) agreed Yes (compared to 22% that said No) and felt that disease prediction using ML should still be used if it is <u>only slightly unfair but better prediction</u> overall compared to current clinical models (e.g., 10% better at predicting disease in women vs men; or disease in one ethnic group vs another ethnic group).

Most (89%) participants offered their own perspective about *fairness* should be determined by. Factors such as historical perspectives and cultural framework concepts like manaakitanga (a Māori term for the process of showing respect, generosity and care for others) were deemed to be important. Fairness that could incorporated impactful solutions for specific ethnic groups, whilst in pursuit of equal health outcomes for all was also offered. Participants highlighted the importance of fairness being representative of groups may require a host of tailored approaches based on a range of specific risk factors. A few participants highlighted the importance of co-design and ensuring that the development of predictive models are constructed according to fair and inclusive processes, thus allowing for vital input from diverse types of stakeholders. There was also a shared recognition, that ML models are not perfect and may have limitations, but that they are worth exploring and developing if there are ultimate benefits and co-benefits to society as a whole. Overall, participants were considerably interested while also holding some concerns about the use of ML models in NZ. There is a desire to carefully explore what potential implications exist before ML models are deployed and implemented with members of the wider public, and most especially amongst vulnerable and high-risk communities that have much higher risk exposure of being excluded from preventable and effective health services.

#### (6) Including community consensus into disease prediction models for fair outcomes

When asked to hold a hui or citizen jury of their community members to debate the issues of:

(1) Official data being given to GPs for improve disease prediction; (2) The trade-offs between under-diagnosis and over-diagnosis – for severe diseases like diabetes and heart disease; and (3) Exploring how fair these systems need to be before being introduced in Aotearoa NZ.

The majority of participants (89%) said that they supported a hui or citizen jury for their respective ethnic communities.

Over half of the participants (67%) positively and 33% were against the use of a Polis system (as tried in Taiwan) for their respective communities in order to achieve an agreed-upon view of using official data to inform disease prediction at the GP level.

One participant supported using an online platform but cautioned against comparing NZ to Taiwan because of significant socio-politico differences between the two countries, which can skew modelling perspectives. Internet access and connectivity also differs amongst different ethnic groups across NZ. Māori and Pasifika communities have lower internet coverage rates than European communities.

Participants mentioned that every community has its own distinctive life challenges, like food security and housing needs. Hosting conversations and allowing people to raise the issues that are important to them and their circumstances may not be beneficial without adequate support to address those issues. Still, overall, participants agreed that engaging with communities this can be highly beneficial, as it allows for two-way communication and builds trust.

Another important matter highlighted by participants was the importance of meeting the community on their own terms and in their own environments. Doing so can demonstrate a credible commitment of building understanding and addressing the concerns experienced within that community. Ultimately, this kind of engagement can help build stronger relationships and lead to more effective solutions, especially regarding new and emerging technologies.

A contrary view that one participant offered about community consultation was that social media platforms may not capture the views of everyone. Even when conducting polls, high response rates may be met with known and unforeseen challenges that in turn can impact the accuracy of results. In this context, researchers and modelling teams would need to work closely with communities in articulating what agreed arrangements were needed in association with cultural safety, language accessibility, ownership and controls of data sources and other complex components. The participants also noted the importance of targeting ethnic groups and ensuring that whatever information is gathered is taken back to the communities involved. By using fair and democratic approaches, researchers and modelling teams can demonstrate mutual respect for a community’s right to self-determination, particularly in relation to data sovereignty frameworks and practices valued by Māori Indigenous communities.

## Discussion and Conclusions

Artificial intelligence (AI) and ML pose enormous potential for improving quality of life. It can also generate significant social, cultural and other unintended risks. The study shows that participants were strongly united in the view that ML models should not create or exacerbate inequities in healthcare due to biased data and unfair algorithms. In NZ, diabetes and CVD are the leading causes of premature death and disease burden and major sources of health inequities for Māori, Pasifika, and Asian populations. An exploration of fairness concepts showed that carefully selected data types must be considered for GP predictive modelling and that trade-offs for obtaining benefits versus generating unnecessary concern produced conflicting opinions. However, participants were comfortable with greater benefits that extended life with slight minor health trade-offs to the least number of people. The participants expressed high acceptability for using ML models but expressed deep concerns about inequity issues and how using these models might affect the most vulnerable communities (such as Māori in middle-ages and above, those living in deprived communities).

Our study addressed several key trade-offs that have been neglected in the ML literature, including the balance between obtaining benefits and generating unnecessary concern, and fairness versus model prediction in real-world scenarios.(Barocas, Guo et al. 2021, Rodolfa, Lamba et al. 2021)

Information from health and data experts showed that they were highly receptive to the use of ML models, indicating a supportive environment in NZ for capitalising on the country’s world-class data linkage and the latest developments in AI. We also identified the potential for obtaining feedback on these models from social platforms in NZ. This supportive environment for ML models suggests that we can avoid the suboptimal case of the AI model for child welfare in the US, where child welfare agencies have turned to predictive analytics without first seeking input from stakeholders. In such cases, the use of these prediction models may perpetuate or exacerbate existing problems in child welfare, according to participant feedback in the US. (Stapleton, Lee et al. 2022)

Concerns regarding inequity issues and how using these models might affect the most vulnerable communities suggests that the development and deployment of the ML models must involve ethics and social issues. Therefore, a community-participatory approach to evaluate the social impacts of these ML algorithms, as the one used in this study, should be implemented before the model is deployed in the community(Crawford and Calo 2016, Krupiy 2020, Vesnic-Alujevic, Nascimento et al. 2020). Data and data-driven tools can be better supported by communities that are most likely to be impacted, thus, pathways to mitigate possible harms of these tools should be approached with serious consideration (Stapleton, Lee et al. 2022). This is of particular importance for NZ with its history of colonialism, Eurocentric paradigms and ethnically diverse communities who would prefer using their own methods and solutions to address concerns that impact them directly, concerns that often affect families intergenerationally.

The strength of our research was that we could perform one-hour in-depth surveys in a consistent format with two interviewers. Our questionnaire was designed in a way that can be potentially used for the development of fair ML models with diverse ethnic communities.

Given the qualitative approach used for the study, a small number of participants may have limited the scope of our findings; however, this was an effective methodology for the in-depth exploration of views, opinions, values and narratives. In addition, this is a recently topical subject within the ML literature, so even well-informed experts who participated in our study found our semi-structured questionnaire format to be challenging, particularly, the sectionof trade-off scenarios. In future, the questionnaire may need to be altered to suit different groups and expertise (eg, technical versus non-technical, clinical versus non-clinical) and be tailored to each specific ML model.

It is important that future ML models are not solely developed based on the opinions of experts and existing literature but that it also incorporates feedback from key informants and the respective ethnic communities that are mostly to be affected. Depending on the nature and emergency of the disease, this feedback loop could operate in real-time, enabling models to improve their predictions based on up-to-date information. For instance, some prediction models can already use real-time data from platforms such as Twitter to improve their accuracy in predicting outbreaks like the flu.(Alkouz, Al Aghbari et al. 2019).

Our results could help inform the development of ML models that consider social impacts in an ethnically diverse society.

## Data Availability

All data are confidential and not available for sharing.

## Statement of Ethics Approval

The study was approved by University of Otago ethics approval processes, reference number HD20/012 and D22/101. There were no patients directly involved in this study.

## Acknowledgments

This work was funded by the Royal Society Te Apārangi. The funders had no role in study design, data collection and analysis, decision to publish, or preparation of the report. We thank Nick Wilson for helping with the survey design and useful comments on an earlier version of this paper. Our special thanks and aroha goes to all the interview participants.

## Authors’ contributions

NN, RR conceived and designed the study. NN, RT designed survey questionnaires. RT, NN performed the interviews with the participants. NN, RT analysed and interpreted the results. NN wrote the first draft, and RR contributed significantly and critically to the writing. NN, RT approved the final draft.

## Appendix A

**Study on making automatic data analysis for disease prediction fair for people of different ethnic groups** (Formal name: “Predicting diabetes complications risks and costs using machine learning with equity analysis”, sub-study)

### KEY INFORMANT INTERVIEW

#### Interview questions

##### I. Existing attitudes

a. Do you think health data in Aotearoa New Zealand (NZ) is currently used to fairly allocate health care to address need, in particular treatment vs prevention?
b. Do you think GPs currently typically have enough individual patient data upon which to base the best health decisions for individual-level care? [additional relevant information that could be predictors for conditions [social determinants of health – nutritional information] eg poverty, food security, benefit entitlement, unemployment]

##### II. Attitudes to data held at the GP level

a. Do you think it is okay for government agencies to give data on individuals (eg, social welfare data) to GPs to improve how <u>GP-level disease prediction</u> models may work – so that all patients (including Māori and Pasifika) get better prevention and disease management?

##### III. Acceptable data to have at the GP level for prediction models

a. What information are you personally comfortable to be provided to GPs from official government sources for health prediction purposes, thinking about your views and those of your community? (Score: 5 Very comfortable, 4 Fairly comfortable, 3 In between comfortable and uncomfortable, 2 Fairly uncomfortable, and 1 Very uncomfortable)

b. Is there any other information you think is acceptable for official sources to provide to GPs for improving the prediction of future health conditions? Please list.
c. Is there any information that you <u>do not want</u> to have used to predict future health conditions for you or your community? Please list.

| Score | Type of information |
| --- | --- |
| **Demographic** |  |
|  | Demographic, including age, sex, ethnicity, education, income, language, province (rural/urban) |
| **Health** |  |
|  | Health history (eg, past heart disease, diabetes, and cancers) and current medications (including from other parts of the NZ health system) |
|  | The amount of alcohol a person typically drinks (eg, if they have revealed this to a health worker in another part of the NZ health system) |
|  | If a person currently smokes tobacco or not, or is an ex-smoker |
|  | How a person typically travels to work, walk or drive etc (eg, from |
|  | Census data) |
|  | A person's diet such as how much sugar a person typically eats (eg, if they have revealed this to a health worker in another part of the NZ health system) |
|  | A person's physical activity levels (eg, if they have revealed this to a health worker in another part of the NZ health system) |
| ***Socio-economic*** |  |
|  | Immigration, such as birth country, time in NZ |
|  | Spending, such as personal or household expenditure |
|  | Employment, occupation, working hours, utilisation, etc... |
| ***Justice*** |  |
|  | Previously being a victim of crime |
| ***All data*** | All administrative data (ie, all information from a person's interactions with the Government) |

#### Scenarios for discussion

##### IV. Trade-offs for obtaining benefits vs generating unnecessary concern

In your view what are the worthwhile trade-offs eg, for Māori/Pasifika patients for serious conditions like heart disease:

a. <u>One</u> patient get slight health benefits from better GP-level prediction – but <u>one</u> is made slightly worse off because of being told they are at risk of an adverse health outcome when this is not actually correct. For example, one extra person with diabetes is put on blood pressure medicine that will help prevent heart attacks but one extra person is put on these medicines unnecessarily. All those who take the medicine gain an average of 6 months of extra life; while the person taking medicine unnecessarily has mild side-effects – some slight extra tiredness and has to visit their GP two times extra per year.
b. <u>Five to 10</u> patients get slight health benefits from better GP-level prediction – but <u>one</u> is made slightly worse off because of being told they are at risk of an adverse health outcome when this is not actually correct.
c. What is your comfortable range of these trade-offs (ie, instead of <u>one or five to 10</u> patients being slightly better off) for <u>one</u> patient slightly worse off?

##### V. Reducing bias in risk prediction models (ie, results of machine learning models)

If disease risk prediction models are not well designed then they can work better for some groups in society rather than others (ie, usually benefiting the groups which are largest in society). What is you view on this:

a. Disease prediction using machine learning should not be used in Aotearoa NZ until it is guaranteed to be fair (performing equally well for all groups and not better for one group than another).
b. Disease prediction using machine learning should still be used if it is <u>only slightly</u> <u>unfair but better prediction</u> overall compared to current clinical models (eg, 10% better at predicting disease in women vs men; or disease in one ethnic group vs another ethnic group).
c. What is fair in your view?

##### VI. Including community consensus into disease prediction models for fair outcomes

a. What are your thoughts on holding a hui or citizen jury of your community members that debated the issues of: (1) Official data being given to GPs for improve disease prediction; (2) The trade-offs between under-diagnosis and over-diagnosis – for severe diseases like diabetes and heart disease; and (3) How fair these systems need to be before being introduced in Aotearoa NZ.
b. In Taiwan there is an online process developed to help society discuss solutions to complex issues. It has been successfully used over 100 times and uses a pro-social media platform. As people interact with the views on the platform the software builds domains of consensus. This means that policy makers can see positions agreed upon by most of the public which to subsequently build policy solutions. What are your views on using a Polis system for your community members to achieve an agreed view on the use of official data to inform disease prediction at the GP level?

